# An Open-Source 3D Printed Simulator for Lumbar Durotomy Repair: A Single Neurosurgery Residency Program’s Experience

**DOI:** 10.1101/2025.04.04.25325265

**Authors:** Robert Winkelman, Ahmed Kashkoush, Gregory Glauser, Swetha Sundar, Richard Schlenk, Ajit Krishnaney, Michael Steinmetz, William Clifton

## Abstract

**Introduction:** Surgical simulation is an appealing adjunct for educating trainees on a variety of spine surgery techniques, especially non-routine skills like lumbar durotomy repair. However, high costs and significant lead time required to obtain surgical simulators restrict their routine use in most training programs. The goal of the present study was to develop and evaluate a low-cost and open-source 3D printed lumbar durotomy repair simulator for resident education.

**Methods:** The design of a pre-existing open-source 3D printed spine simulator was modified for a durotomy repair simulation. The simulator was then printed using acrylonitrile butadiene styrene (ABS) filament on a Bambu Lab P1S printer. Current neurosurgical residents from a single institution were recruited to participate in a one hour lab session in which they were tasked with repairing a standardized 1.5 centimeter durotomy. Subjects were surveyed before and after their simulation experience.

**Results:** Seven simulators were produced taking on average 12.2 hours to print and cost approximately $9.51 each once fully assembled. Of the 14 subjects recruited, only 7 (50%) reported prior experience with lumbar durotomy repair. All subjects were able to successfully complete the simulated task. Exit survey results demonstrated that all subjects (100%) agreed the simulation was useful and expressed interest in participating in future simulation experiences.

**Conclusions:** Our study demonstrated a 3D printed simulator for lumbar durotomy repair could be produced at low cost and was highly valued by neurosurgical trainees. Given our simulator’s low cost and open-source format, we believe it is highly accessible to most, if not all, residency programs, and has the potential to help expedite mastery of lumbar durotomy repair.

## Introduction

Providing trainees opportunities to perform lumbar durotomy repair in the operating room is a common dilemma for spine surgery faculty. While mentored practice is essential for trainees to develop into competent spine surgeons, inadequate durotomy repair can lead to postoperative complications and negatively impact patient outcomes. Although graduated autonomy and years of experience are critical for achieving surgical mastery, surgical simulation offers a promising adjunct for trainee education in durotomy repair.

Prior studies across multiple surgical specialties have demonstrated effective surgical simulators can accelerate trainee surgical skill acquisition and performance in the operating room.^1^ Unfortunately, high costs and often significant lead times needed to obtain commercial surgical simulators often restrict their routine use in many training programs.

Several non-commercial durotomy repair simulators have been described, ranging from sophisticated models using perfused cadaveric specimens to more simplistic models using basic materials. ^2,3^ While preliminary results suggest these simulators may enhance trainee mastery of durotomy repairs, challenges related to the acquisition, preparation, and storage of cadaveric tissue and lack of relevant surgical anatomy in more simplistic simulators likely limit their incorporation into many residency curricula. Simulators developed with three dimensional (3D) printers may represent a potential alternative to previously reported durotomy repair simulators as they can be produced at low cost and offer more realistic surgical anatomy compared to more basic simulators. 3D printed simulators have been successfully used to educate residents for the placement of freehand lumbar pedicle screws and are available in open-source format.^4^ However, open-source 3D printed simulators for durotomy repair are currently lacking.

The goal of the present study is to develop and assess the use of a 3D printed lumbar durotomy repair simulator for neurosurgery resident education at a single institution. This manuscript will detail the methods used to develop and produce an open-source 3D printed simulator for lumbar durotomy repair. The manuscript will also present the results of a pilot study where the simulator was used as a part of a neurosurgery resident skills lab.

## Methods

### Development and assembly of 3D printed model

The model was developed by modifying an open-source 3D printed lumbar spine model “The Spine Box.”^4^ The standard tessellation language (STL) file was modified by adding a luer lock attachment and nozzle to spine box to accommodate attachment of simulated thecal sac in spinal canal.^5^ Modifications to the model’s STL were made using the open-source software Blender.^6^ The modified STL file of the model is publicly available for download.^7^ The model was then printed using acrylonitrile butadiene styrene (ABS) filament on a Bambu Lab P1S printer using the standard ABS print profile on BambuStudio.^8^ Once printed, a laminectomy was performed at L3 using a combination of leksell and kerrison ronguers. To create a simulated thecal sac, a 5/8th x 12 inch silicone penrose drain was tied off at one end and trimmed to approximately 7 inches in length. The open end of the penrose was passed through the spinal canal and connected to the opposite end of the luer lock nozzle with a 2-0 silk tie. A 500 cc normal saline bag was connected to IV tubing and attached to the luer-lock on the outside of the 3D printed portion of the simulator.

Silicone sealant was used to reinforce the IV tubing and luer-lock attachment to prevent inadvertent leaking of saline at the connection site. The saline bag was used to fill the attached simulated thecal sac. Lastly, a seven by seven inch piece of two-inch thick upholstery foam was cut and placed over the spine model in the modified Spine Box to simulate skin/paraspinal soft tissue and recreate the “working in a hole” experience of spine surgery. An example of the assembled simulator can be seen in Figure 1A. **Subjects** Subjects were voluntarily recruited from our institution’s neurosurgery residency program to participate in a one-hour simulation lab using the aforementioned 3D printed simulators for durotomy repair. Residents were electronically surveyed before their participation in the simulator lab and asked to self-report their prior experience with durotomy repair as well as four other common neurosurgical procedures (lumbar laminectomy, freehand lumbar pedicle screw placement, hemicraniectomy, and suboccipital craniectomy). In addition to the number of prior experiences, the survey also asked subjects on the type of prior experiences they had performing these procedures (i.e. in a simulated setting, in cadaveric specimen, or in the operating room). The study procedures were reviewed and approved by our institutional review board.

**Figure 1A +B.**
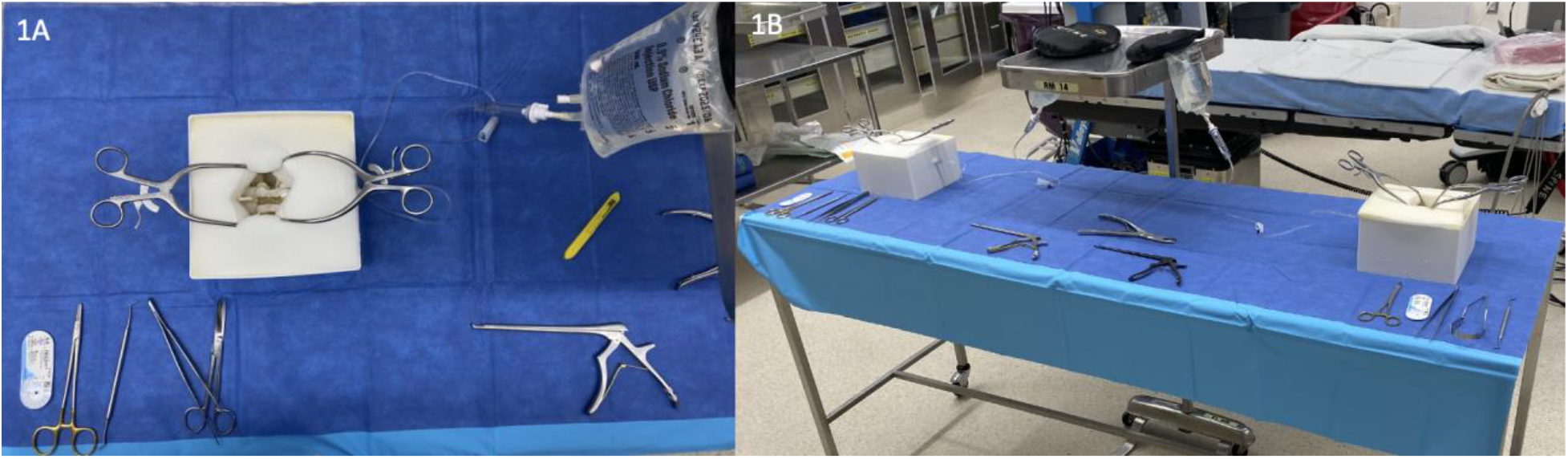
A: View of assembled simulator. B: View of example set-up in operating room with two simulators on single stainless steel stable with mayo stand in middle suspending IV fluid backs to height of approximately 30 cm above the durotomy defect.

### Simulation Lab

The lab was completed during a one-hour morning time slot which is normally reserved for resident education. The lab was held in a vacant operating room configured with four stainless steel tables with two simulators on each table. Each station was provided with a needle driver, fine-toothed forceps, 6-0 prolene suture, Metzenbaum scissors, two slightly angled cerebellar retractors, and kerrison ronguers. Prior to the start of the lab, a 1.5 cm midline incision was made in the simulated thecal sac using a #15 blade to simulate a durotomy. A mayo stand was positioned in the middle of the table between the two simulators and raised to a height where the 500 cc normal saline IV bag would hang approximately 30 cm above the durotomy defect. Set-up of an example lab station can be seen in Figure 1B.

Subjects were split into pairs (i.e. one junior with one senior resident) and instructed to perform a watertight repair of the simulated dural defect using 6-0 prolene suture. A simulated valsalva using the elevated IV bag would be used to assess the integrity of the repair. All subjects were encouraged to bring loupes, headlight, and surgical gloves.

Subjects were assessed for the ability to complete the simulated procedure by the study investigators. Time to complete the simulated procedure was also recorded.

### Post Survey

After completion of the simulation experience, subjects were electronically surveyed again to assess their simulation experience. Specifically, subjects were asked to respond on a five-point Likert scale to questions on the following five topics: 1) if they felt they had sufficient time to complete the simulated procedure 2) the quality of simulated dural tissue, 3) the utility of simulation for skills practice, 4) their interest in participating in future simulations, and 5) their interest in using similar simulators for possible independent study. All study data were aggregated and analyzed in R and RStudio.^9,10^ Analysis was limited to descriptive statistics using percentages for categorical variables and averages for continuous variables.

## Results

### Production of 3D printed simulators

Seven lumbar spine simulators were printed using 350 g of ABS filament per model. Each print took approximately 12.2 hours for a total of 85.4 hours (3.6 days) of total print time. No print failures encountered.

After printing, each model took approximately ten minutes to assemble for a total assembly time of approximately 1.25 hours for all seven models. After accounting for all materials used to assemble each simulator, the estimated total cost for each simulator was approximately $9.51 (Table 1).

**Table 1.**
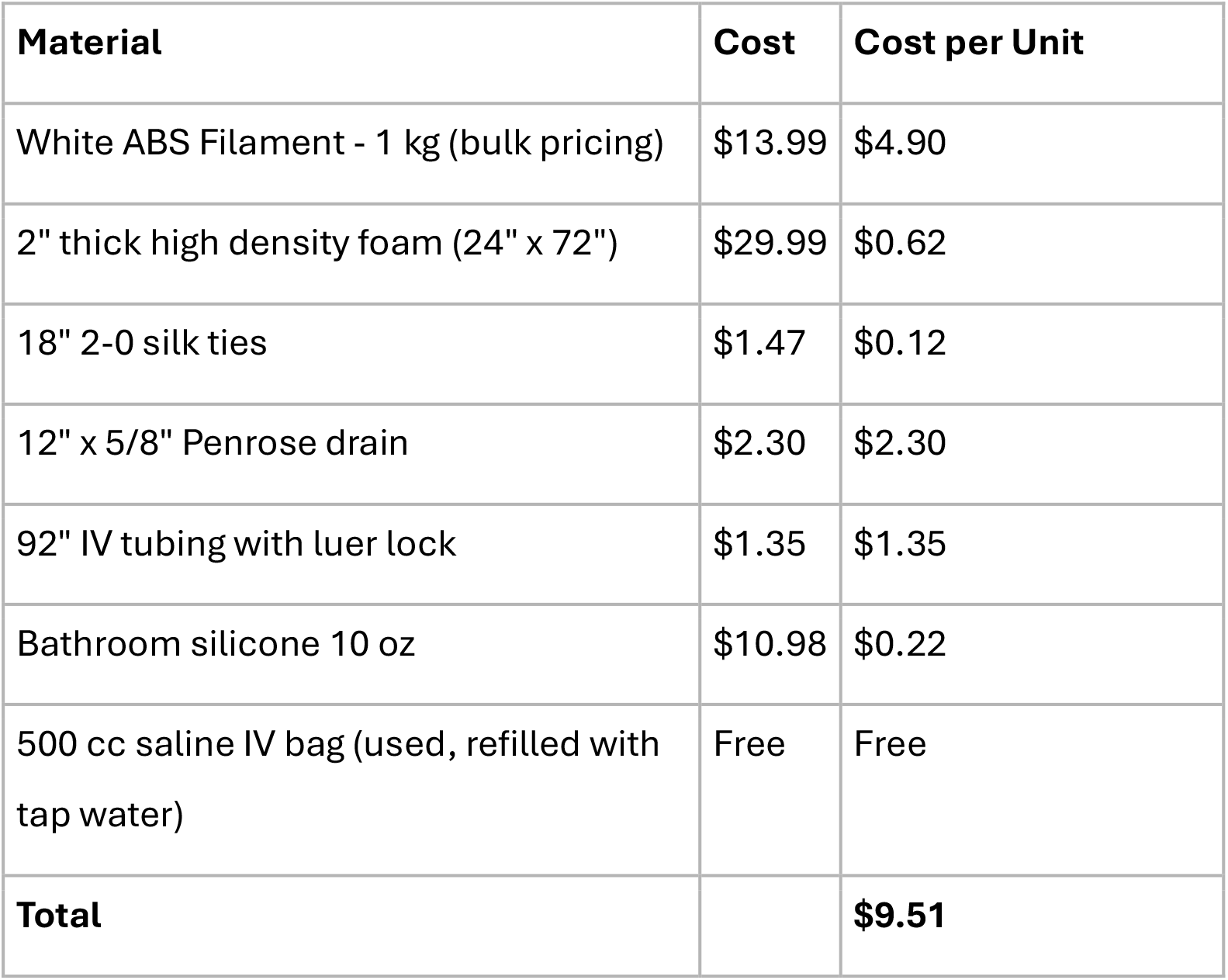
List of materials and estimated cost.

| Material | Cost | Cost per Unit |
| --- | --- | --- |
| White ABS Filament - 1 kg (bulk pricing) | \$13.99 | \$4.90 |
| 2" thick high density foam (24" x 72") | \$29.99 | \$0.62 |
| 18" 2-0 silk ties | \$1.47 | \$0.12 |
| 12" x 5/8" Penrose drain | \$2.30 | \$2.30 |
| 92" IV tubing with luer lock | \$1.35 | \$1.35 |
| Bathroom silicone 10 oz | \$10.98 | \$0.22 |
| 500 cc saline IV bag (used, refilled with tap water) | Free | Free |
| **Total** | | **\$9.51** |

### Resident Survey and Simulation Lab Experience

Fourteen subjects were recruited to participate in the simulation experience. Subject by post-graduate year of training can be seen in Table 2.

**Table 2.** Subject year of training.

| Subject Year of Training | Count |
| --- | --- |
| PGY-1 | 1 |
| PGY-2 | 3 |
| PGY-3 | 2 |
| PGY-4 | 3 |
| PGY-5 | 2 |
| PGY-6 | 3 |
| PGY-7 | 0 |
| Total | 14 |

Subject self-reported prior experience with five common neurosurgical operating room procedures as well as setting where they had the opportunity to practice the procedures are presented in Figure 2.

**Figure 2.**
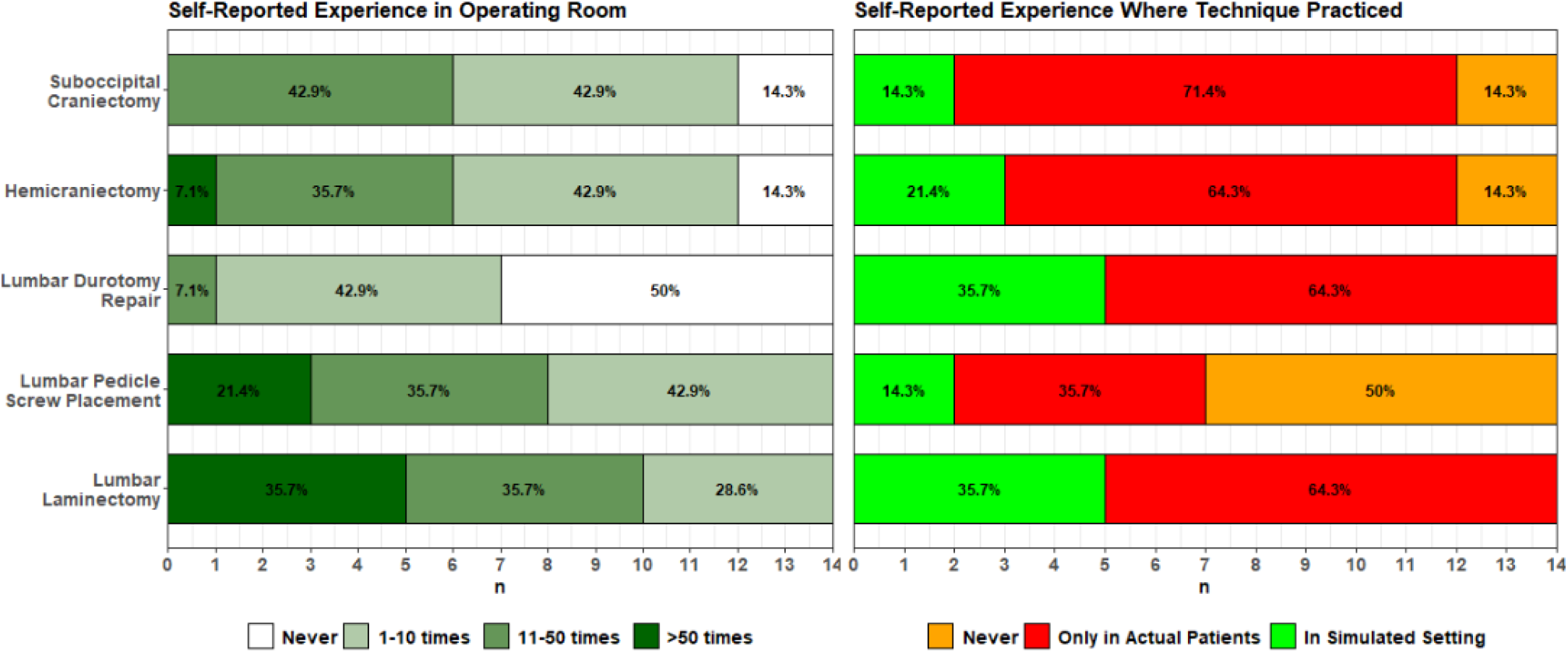
Distribution of subjects’ self-reported prior experience and setting of prior experience for five common neurosurgical procedures

In regard to their experience with the five common operating room procedures, subjects reported the least experience with lumbar spine durotomy closure, with 50% (n = 7) reporting no prior experience with durotomy closure. Of note, only two subjects with prior durotomy closure experience had reported prior experience practicing durotomy closure in a simulated setting while the other five subjects reported their only durotomy closure experience had been in actual patients.

During the laboratory session, all 14 residents were able to complete the simulated procedure of watertight closure during the 1-hour period. The time to complete the repair took an average of 15.8 minutes per subject.

Figure 3 shows the results from post-simulation experience survey completed by 13 of 14 participating subjects. All subjects agreed that they had sufficient time to complete the simulated procedure. Subjects also agreed they would be interested in participating in further simulations as well as the opportunity to use the simulators for independent skills practice. The majority of subjects also agreed the simulated dural tissue felt comparable to real dural tissue.

**Figure 3.**
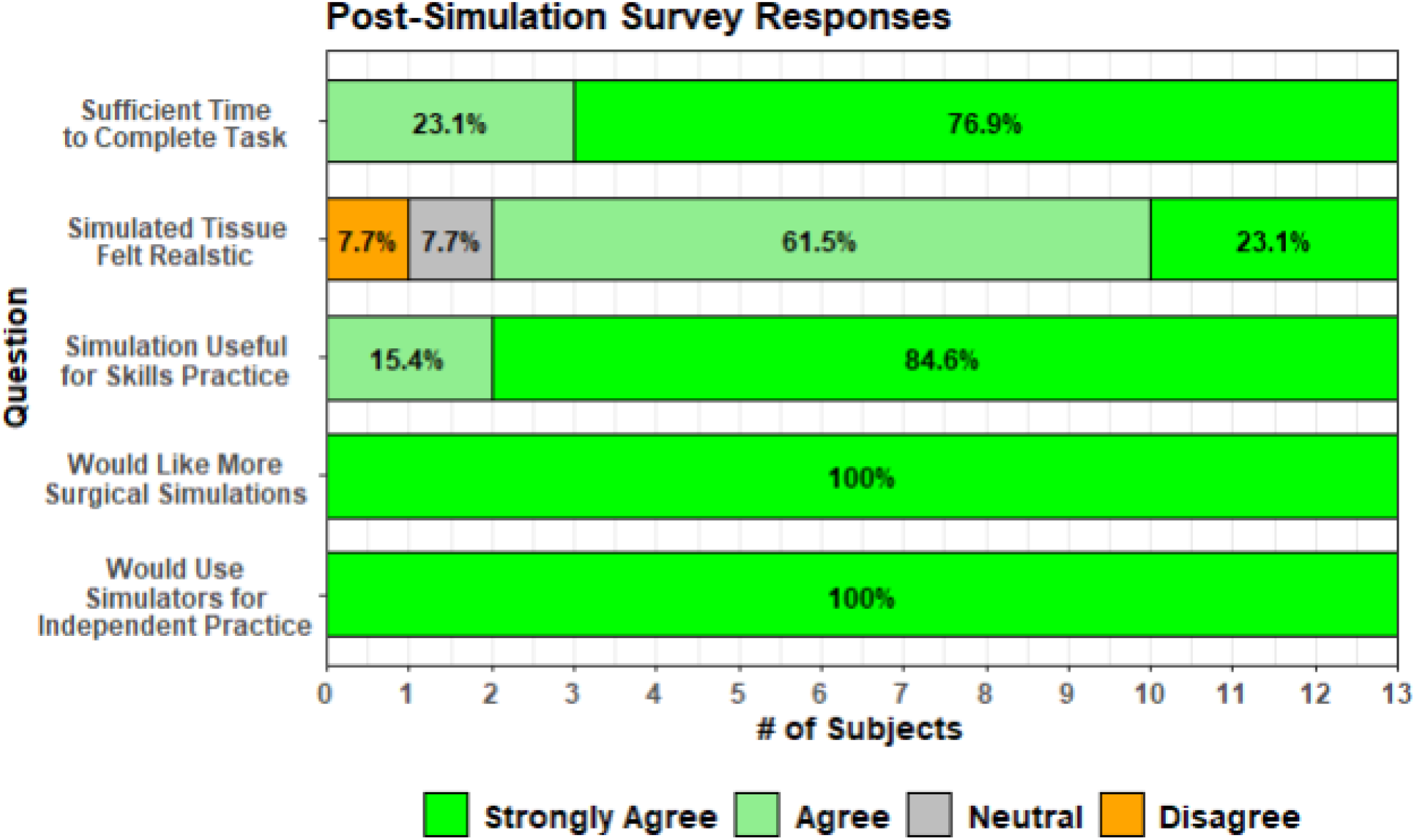
Distribution of subjects’ responses to post-simulation survey

## Discussion

Given the challenges associated with providing trainees opportunities to practice durotomy repair and the current lack of accessible surgical simulators, we developed an open-source 3D printed simulator for durotomy repair and evaluated a single neurosurgery residency’s experience with it during a one-hour simulation lab. The simulators were produced at low cost and required little time to assemble. Pre-simulation survey results indicated that subjects had relatively less experience performing durotomy repair compared to other common operating room procedures. All subjects successfully completed the simulation. Post-simulation survey results indicated subjects found the experience valuable and expressed interest in participating in future simulations.

Our findings suggest that the use of 3D printed simulators for lumbar durotomy repair practice may help to address a potential gap in neurosurgery resident education, as subjects in our study reported less experience with durotomy repair compared to other common operating rooms procedures. There are likely several factors that contribute to trainees’ limited opportunities to practice lumbar durotomy repair. First, lumbar durotomies are likely less frequent than other procedures and often represent an unplanned complication of lumbar spine surgeries. Second, the technical skills needed to perform a lumbar durotomy repair are likely higher than other common procedures and thus may be more appropriately reserved for more senior residents, fellows or attending surgeons. Lastly, attending surgeons may limit trainee involvement in this portion of the case due to concerns about potential complications from inadequate repairs. Regardless of the specific reason behind the limited opportunities for trainees to complete durotomy repair, low-cost 3D printed simulators can provide trainees more opportunities to practice the microsurgical skills needed for durotomy repair. Prior literature has demonstrated that repeated practice opportunities with surgical simulators may correlate with improved trainee performance as it allows them the opportunity to develop fine motor control and dexterity with instruments.^11^ Surgical simulation also enables learners to rehearse less common scenarios in a no-risk setting. For example, although the durotomy repaired in the present study was a simple midline durotomy, the simulated durotomy could be modified to a more challenging location (i.e. a durotomy in a lateral or ventral location) to allow more senior learners opportunities to practice more non-routine repairs. Given the limited time needed to assemble the simulators and perform the durotomy repair lab, the described durotomy repair simulation could be an appealing adjunct to other neurosurgery residency curriculums.

Importantly, the surgical simulator in the present study is produced at low cost and access to the files needed to produce it is freely available to the public in an open-source format. Other 3D printed simulators which could be used for lumbar durotomy repair practice have been published, but the details on their production and assembly have not been shared for free public use.^12,13^ Our durotomy repair simulator is also a modification of another open-source simulator (SpineBox)^4^ and thus effectively demonstrates how democratizing access to 3D printed simulators can enable others to iterate and enhance existing simulators to address specific educational objectives. Falling costs and increased ease of use of 3D printing technology are also reducing financial and technical barriers which may have previously prevented the more widespread adoption of 3D printed simulators. For example, the printer used to produce the model in the present study was purchased for under $800, which is approximately one quarter of the cost of the printer used to produce the initial SpineBox described by Clifton et al.^4^ Additionally, the simulators in the present study were printed on standard print settings (with no need for special printer calibration or fine tuning) and no print failures were encountered during the production of models for this study. Thus, although some commercially available simulators may provide higher quality simulated tissues, the production of open-source simulators such as the one in the present study are likely more accessible and could be more routinely used even in the most financially constrained graduate medical education programs. Given the low upfront capital requirement, low technical barrier and low cost per model, it is conceivable that the financial and technological resources needed to create such simulators are within the reach of most, if not all, neurosurgery training programs.

Although our study demonstrates promising results for the use of a free and open-source 3D printed simulator for lumbar durotomy repair, there are several important limitations to our findings. Our study showed that trainees successfully completed the simulated procedure and reported a positive experience; however, it is unclear whether this translates to improved performance in the operating room. Given prior studies have shown simulators can enhance surgical skill acquisition and performance, it is plausible that our 3D printed durotomy repair simulator may also enhance skill acquisition and surgical performance. However, further research is needed to validate these potential benefits. Additionally, although most subjects reported satisfaction with the simulation experience and simulated tissues, we acknowledge our simulator is low fidelity compared to other durotomy repair simulators. Enhancements such as alternative dural analogs (e.g. bovine pericardium), lumbar nerve roots which could herniate through the dural defect, and simulated epidural bleeding after durotomy could also provide trainees with a more realistic experience of actual durotomy repair. However, prior work has shown that even simplistic simulators can provide comparable skill improvement to skill improvement seen in more sophisticated models.^14,15^ Therefore, while such simulator enhancements could improve realism of the simulation, their adoption must be balanced against the potential increased time and cost of production, which could in turn make the simulator impractical for more widespread or routine use.

## Conclusion

Our study demonstrated the successful development and utilization of 3D printed surgical simulators for durotomy repair at a single US residency training program. Pre- and post-simulation survey results demonstrated that study subjects had relatively limited experience with lumbar durotomy repair and these subjects found the one-hour session with the 3D printed simulators beneficial for lumbar durotomy repair practice. Thus, by providing residents more opportunities for microsurgical skills practice, our simulator can likely help expedite resident mastery of lumbar spine durotomy repair. Additionally, given its low cost, relative ease of production and open-source format, this simulator is currently accessible to most, if not all, neurosurgery training programs.

## Data Availability

All data produced in the present study are available upon reasonable request to the authors

## Notes

### Competing Interest Statement

The authors have declared no competing interest.

### Funding Statement

This study was partially funded by Cleveland Clinic Foundation Catalyst Grant.

### Author Declarations

Ethics committee/IRB of Cleveland Clinic Foundation gave ethical approval for this work

